# Effect of Role of Robotic Assistance on Upper-limb Sensorimotor Recovery: A Systematic Review and Meta-Analysis

**DOI:** 10.1101/2025.04.24.25326375

**Authors:** Ritwika Choudhuri, John Solomon, Aravind Nehrujee, S. Sujatha, Sivakumar Balasubramanian

## Abstract

**Background:** Robot-assisted training is increasingly being adopted to deliver high-intensity, repetitive, and task-specific treatments with real-time feedback to augment recovery. However, the causal role of robotic assistance in sensorimotor recovery, an underlying core mechanistic feature, remains unclear.

**Objective:** This study systematically reviewed randomized controlled trials (RCTs) on upper-limb rehabilitation that isolated the effect of robotic assistance by comparing similar interventions differing only in presence, level, or mode of robotic assistance.

**Method:** Three electronic bibliographic databases-PubMed, Scopus and Google Scholar were searched. Twelve RCTs were included in the qualitative synthesis, and data from eight studies (all involving stroke survivors) were meta-analysed with the Fugl-Meyer Assessment score (measure of impairment) as the primary outcome. The main analysis was done to assess the overall effect of robotic assistance, regardless of disease stage, assistance level and targeted limb area, whereas subgroup analyses were conducted to evaluate the impact of robot-assisted therapy on chronic stroke survivors and to compare therapies focused on the shoulder-elbow versus the hand (wrist/fingers).

**Results:** The main analyses yielded a significant positive effect size (0.462) favouring robotic assistance in impairment recovery, however with a significant heterogeneity between the studies. Subgroup analyses suggested potential benefits in chronic phase (moderate significant effect size 0.588), and similar therapy gain for shoulder-elbow and hand-specific training.

**Conclusion:** Limited available RCTs with small sample sizes and moderate heterogeneity prevented any firm conclusion to be drawn, and highlighted the need for systematic, controlled studies with large sample sizes to better understand the direct role (mechanistic feature) of robotic assistance in sensorimotor recovery.

## Introduction

Robot-assisted upper-limb neurorehabilitation has been an active area of research for over 25 years^1^. Various robotic devices^2,3^ have been designed and developed to facilitate rehabilitation, targeting specific joint movements (e.g., shoulder, elbow, wrist), coordinated arm movements (e.g., shoulder-elbow interactions), hand movements, and whole upper-limb movements. A recent review^4^ reported that robot-assisted upper limb rehabilitation therapies can significantly improve motor impairment, albeit small, with early intervention (<3 months post-stroke) leading to better upper limb capacity. Robotic rehabilitation systems provide real-time kinematic and force feedback, enabling interactive gamified therapy sessions that engage patients more effectively than traditional rehabilitation methods. Unlike sensor-based systems, robots can physically interact with human subjects by applying controlled forces and moments to assist or resist movements^5,6^. The primary mode of physical interaction in robot-assisted therapy involves movement or task assistance, wherein the robot exerts controlled forces on the patient’s limb to facilitate and complete movements^7^. Robotic assistance augments voluntary effort to make larger movements by moving the limb outside its active range of motion and helps in performing faster movements than is possible voluntarily, thus, boosts patients’ motivation as well^8^. Assist-as-needed control strategies, in which robotic assistance is dynamically adjusted based on the patient’s effort, has been shown to enhance motor learning and promote voluntary engagement, which is critical for neuroplasticity and functional recovery^9,10,11^.

Given that robotic assistance is a core component of robot-assisted therapy, it is natural to ask the following fundamental causal question: does robotic assistance play a role in driving sensorimotor improvements? The answer to this question holds significant practical value. Clinically, it can inform appropriate training protocols with robots, and from the engineering perspective, it can guide the appropriate design of robots and their control strategies for neurorehabilitation. However, addressing the posed question using traditional randomized controlled trials that compare robot-assisted therapy with conventional therapy is challenging, because these two interventions differ in several important ways, beyond robotic assistance. The key differences include the following: a) type of movements trained: conventional therapy often involves therapist-guided exercises with variability, while robotic therapy emphasizes repetitive, highly structured movements; b) feedback mechanisms: robots provide real-time haptic and visual feedback, whereas conventional therapy relies on therapist feedback (mostly verbal); c) training intensity and dose: robotic therapy offers high-intensity and high-dose training, which is often different from conventional therapy; and d) the amount and quality of social interactions vary between the two approaches^3,7^. Isolating the causal role of robotic assistance in sensorimotor improvements requires controlling for these confounding variables. Ideally, comparisons must be made between interventions that differ only in the presence (assisted or unassisted), level (high or low assistance), or mode (fixed assistance or assist-as-needed) of robotic assistance, while other confounding factors are identical or comparable between the groups, such as robots used for training, movement tasks, feedback, therapy intensity/dose, and therapist supervision. Such studies in the current literature could be synthesized and combined to answer the question of the role of robotic assistance in sensorimotor improvements. Currently, there is no systematic review or meta-analysis which investigates the role of robotic assistance on sensorimotor recovery of upper-limb, therefore, this study aimed to address this research gap. It provides the first qualitative and quantitative summary of the effects of robotic assistance on upper limb sensorimotor impairments and activities from randomized controlled trials (RCTs) with groups differing only in terms of the presence, level or mode of robotic assistance.

## Methods

The study extraction was conducted using three electronic bibliographic databases: PubMed, Scopus, and Google Scholar up to July 2024. Various keyword combinations synonymous with the primary Medical Subject Heading (MeSH) terms – robot, rehabilitation, and upper-limb – were employed to define the search strategy. In each database, the search was conducted using a boolean logic rule for the keywords such as “robot-assisted OR robotic OR robot-based OR end-effector OR exoskeleton” AND “rehabilitation OR neurorehabilitation OR therapy OR training” AND “upper limb OR upper extremity OR arm OR forearm OR shoulder OR elbow OR hand OR finger” NOT “review” etc. (detailed keywords and search formulas are provided in the supplementary document, Section-I). These terms were searched for in title, abstract and keywords of the literature. References of the matched articles were imported into the Covidence software^12^ for de-duplication and screening. Based on the selection criteria outlined in Table 1, the studies were screened by reviewing their titles, abstracts, and, where necessary, the full texts.

**Table 1:** List of inclusion and exclusion criteria for determining the eligibility of randomized controlled trials on upper limb robotic therapy to be shortlisted for the systematic review.

|  | Inclusion Criteria | Exclusion Criteria |
| --- | --- | --- |
| **Population** | • People having upper-limb impairment due to neurological disorders such as stroke, spinal cord injury, multiple sclerosis, traumatic brain injury<br>• Participants’ age $\geq 18$ years | • People having upper-limb impairment due to musculoskeletal disorders<br>• Participants’ age $< 18$ years |
| **Intervention** | • Robot-assisted therapy for upper-limb neurorehabilitation (active assistance, assist-as-needed, high assistance, assistance, assistance contingent on movement intention, progressive arm weight support, deficit force-field, negative viscosity force)<br>• Both experimental and control groups in a study were exposed to the same robotic device for therapy<br>• Intensity and dosage of the therapy were same between the groups<br>• Nature and schedule of the feedback were the same between the groups | • Robot-assisted therapy for hip, knee, lower-limb, balance and gait<br>• Error-augmenting robotic therapy<br>• Subjective factors such as active participation and motivation of the patients were implicitly or explicitly manipulated and were different between the groups |
| **Comparator** | • Unassisted, predefined amount of assistance, low assistance, assistance non-contingent on | • End-effector versus exoskeleton, or, proximal versus distal robots, or two |
|  | movement intention, no or full arm weight support, and null force field | same purpose robots with different features or of different brand<br>• Conventional therapy versus robot-assisted therapy<br>• Low versus high intensity robotic training |
| **Outcomes** | • Measure of impairment<br>• Measure of activity and/or performance |  |
| **Study Design** | • Randomized controlled trials (RCTs)<br>• Published in peer-reviewed journals | • Case study or abstract only<br>• Review study |
| **Language** | • English |  |

Two reviewers (RC and SB) independently screened the references and resolved the conflicts through discussion, resulting in twenty studies being shortlisted. Among these, two studies on active versus complete passive assistance^13,14^, and three studies on assistive versus resistive robotic therapy^15,16,17^ were excluded because the active participation of the subjects between the control and experimental groups appeared to be implicitly different in these studies. Moreover, one study on high versus low robotic assistance^18^ which apparently qualified the eligibility criteria got further eliminated as it was a retrospective analysis of data from an RCT^19^, wherein, the control group received self-guided therapy, and the assignment of level of robotic assistance to the individuals in the experimental group was not randomized. Another study^20^ that satisfied the eligibility criteria was also excluded, because it reported preliminary data from another study^21^ which has already been included in this review. One additional relevant study^22^ was removed because it appeared to be part of a larger trial for which no record could be found. Hence due to the lack of clarity on the study’s randomization and blinding procedures, it was not included in the core synthesis of this review. However, the main meta-analysis was repeated including the data from this study^22^ and the corresponding result is reported in Section IV of the supplementary document. In total, twelve studies were finally involved for this systematic review, and data from eight studies (all stroke survivors) were used for the meta-analysis. A flow diagram of the study selection process is shown in Figure 1.

**Figure 1:**
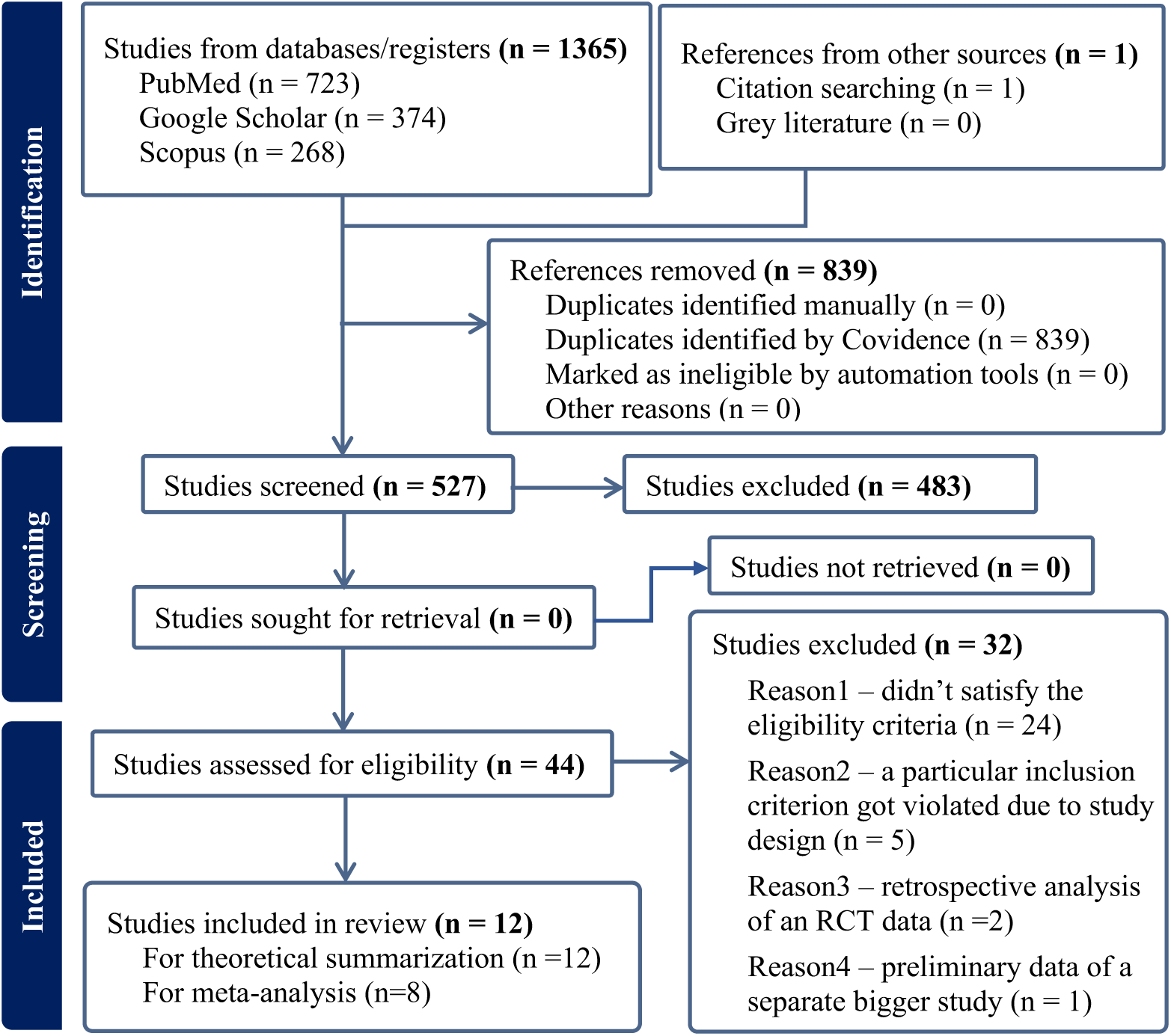
PRISMA flow diagram depicting the steps of the study selection process.

The selected publications were reviewed, and the following data were extracted for qualitative summary and meta-analysis: number of subjects, time from stroke onset, robot employed, experimental protocol, therapy intensity, outcome measures, and pre-post intervention impairment score. Details of this information are presented in Tables 2, 3 of this document and Section-II of supplementary document. The risk of bias (ROB) analysis of all twelve studies was performed manually for the corresponding signalling questions based on individual judgment of author RC upon reading each study literature. The associated document link and the references are provided in Section-III of the supplementary document. The ‘traffic light’ plot for visualizing the ROB report was generated using the open-source software named ‘robvis’^23^ and has been reported in the result section. Analysis of publication bias was also performed for the eight studies which were involved in the meta-analysis; the corresponding details are mentioned in Section-III, and the finding is reported in Section-IV of the supplementary document.

**Table 2:** Relevant information extracted from the studies included in this systematic review. The table summarises the sample detail, study protocol, intervention, and outcome measures.

| Studies | Disease stage and severity | Exp. Group | Control Group | Device employed and Therapy duration | Tasks performed during robot-assisted therapy | Outcome measures | Usual therapy? |
| --- | --- | --- | --- | --- | --- | --- | --- |
| <sup>29</sup>Takahashi et al., 2008 | Chronic stroke<br>Severity- moderate | Active assistance for all sessions (N=7) | Half of the sessions- unassisted but connected to the device, another half of the sessions- active assistance (N=6) | Hand Wrist Assistive Rehabilitation Device ('HWARD')<br>15 sessions, (1.5 hr / session) over 3 weeks | Hand in grasp and release movement (wrist and finger flexion -extension) | ARAT, FMA, BBT, ROM, MAS, NHPT, EMG, fMRI | Not mentioned |
| <sup>35</sup>Ellis et al., 2009 | Chronic stroke (greater than 6 months post stroke)<br>Severity- moderate to severe | Robotic training (no assistance) with progressive arm-weight support (N=7) | Robotic training (no assistance) with full arm-weight support (N=7) | Haptic MASTER<br>3 sessions, in a week over 8 weeks | Reaching movement to targets in 5 directions (6 torque conditions) involving shoulder abduction-adduction, shoulder flexion and elbow flexion-extension | Work area and isometric strength | Not mentioned |
| <sup>34</sup>Huang and Patton 2013 | Chronic stroke (greater than 1 year to less than 9 years post stroke)<br>Severity- severe | Two equal subgroups (cross over design)- Null force, negative viscous force and negative viscous force with inertia (N=20) | Null force field (N=10) | Planar force-feedback device<br>3 sessions, each of 2hr 15 min | Any movement of own choice using a variety of directions, speed and positions within a rectangular workspace (exploratory movement) | Movement quality and performance accuracy in terms of trajectory path, speed and acceleration | Not mentioned |
| <sup>28</sup>Ramos - Murguialday et al., 2013 | Chronic stroke (greater than 6 months post stroke)<br>Severity- severe | Active assistance contingent on movement intension (N=16) | Active assistance non-contingent on movement intension (N=14) | Hand Orthosis with BCI (EEG based)<br>20 sessions; (5 sessions / week); total 4 weeks | Reach and grasp task involving upper arm (shoulder-elbow flexion and extension) movement to reach forward and finger flexion-extension (hand) to grasp | FAM, MAL, GAS, EMG, fMRI | Yes |
| <sup>31</sup>Susanto et al., 2015 | Chronic stroke (6 to 24 months post stroke)<br>Severity- moderate | Robot assisted (N=9) | Unassisted (N=10) | A custom-designed hand exoskeleton robot; 5 individual digits powered by 5 linear actuators<br>3-5 times, 1 hour session per week; 20 sessions in 5 weeks | Finger flexion and extension (hand grasping-opening, three-finger and two-finger pinching-opening) | ARAT, WFMT, FMA and finger individuation index | Not clear |
| <sup>36</sup>Frolov et al., 2017 | Subacute stroke (1-6 months post stroke) and Chronic stroke (greater than 6 months post stroke)<br>Severity- severe | Assistance dependent on motor imagery-based EEG activity (N=55) | Assistance non-dependent on motor imagery-based EEG activity; sham BCI (N=19) | A custom-designed BCI-Controlled hand Exoskeleton<br>10 sessions; each lasting up to 40 min; over 2-3 weeks | Three mental tasks- motor relaxation, motor imagery of paretic hand opening, and motor imagery of non-paretic hand opening.<br>BCI active- the task-imagination specific brain activity drove the exoskeleton attached to paretic hand<br>BCI passive- random actuation of the exoskeleton | ARAT and FMA | Yes |
| <sup>33</sup>Frullo et al., 2017 | Chronic incomplete spinal cord injury (SCI)<br>Severity- moderate to severe | Assist as assisted (N=7) | Self-triggered (partially passive assistance) (N=7) | MAHI EXO-II (upper extremity exoskeleton)<br>10 sessions (90 min each), 1-3 sessions per week, all sessions over 3-4 weeks | Point to point movement from a centre target to peripheral targets involving elbow flexion-extension and wrist radial-ulnar deviation | ARAT, MAS, grip-pinch strength, GRASSP and movement kinematics | Not mentioned |
| <sup>32</sup>Rowe et al., 2017 | Chronic stroke (at least 6 months post stroke)<br>Severity- moderate | High assistance; caused average 82% success rate (N=15) | Low assistance; caused average 55% success rate (N=15) | FINGER exoskeleton<br>1 hour / session; 3 times in a week for 3 weeks; total 9 sessions | Index and middle finger flexion-extension | BBT, FMA, ARAT, MAS, MAL, IMI | Not mentioned |
| <sup>22</sup>Wright et al., 2018 | Chronic stroke (greater than 8 months post stroke)<br>Severity - moderate to severe | Assist-as-needed; deficit force field (N=11) | Unassisted; null force field (N=11) | A custom-designed planar robotic manipulandum<br>3 days / week (2-7 sessions) for 2 weeks; each session of 33-35 min including 1-3 min rest | Planar shoulder-elbow movement (exploratory movement task) | FMA, ARAT, MAS, CMAS-S and elbow ROM | Not mentioned |
| <sup>27</sup>Cho and Song 2019 | Chronic stroke (at least 6 months post stroke)<br>Severity- mild | Assist-as-needed force (N=19) | Guidance force; constant fixed assistance but not completely passive (N=19) | Whole arm manipulandum (WAM)<br>40 min per day, 3 days in a week for 6 weeks | Reaching movement toward targets in 3D space in 6 directions involving shoulder and elbow movements | FMA, ARAT, BBT and movement velocity | Not mentioned |
| <sup>30</sup>Solaro et al., 2020 | Multiple Sclerosis | Haptic- robot assisted (N=19) | Sensorimotor - visual control- no force applied by the robot (N=17) | Planar robotic manipulandum<br>40 min per session, 2 sessions per week for 6 weeks | Fast and accurate reaching movements in 6 different directions | 9HPT and ARAT | Not mentioned |
| <sup>24</sup>Park et al., 2020 | Subacute and Chronic stroke (at least > 3 months)<br>Severity- moderate | Active robotic assistance with gravity compensation (N=10) | Unassisted; mechanical support with gravity compensation by the robot; no actuation (N=9) | Armeo Power (for active assistance group) and Armeo Spring (for passive group)<br>30 min session / day, 5 days in a week for 4 weeks (total 20 sessions) | Virtual reality games involving shoulder, elbow and forearm movements | WFMT, FMA, SIS, and movement kinematics | Yes |

*BCI- Brain Computer Interface, FMA-Fugl-Meyer Assessment, CMSA- Chedoke-McMaster Stroke Assessment, MSS- Motor Status Scale), BBT- Box and Block Test, NHPT- Nine Hole and Peg Test, WFMT- Wolf Motor Function Test, FIM- Functional Independence Measure, ARAT- Action Research Arm Test, MAL- Motor Activity Log, MAS- Modified Ashworth Scale and FAS- Functional Assessment Scale, GRASSP- Graded Redefined Assessment of Strength, Sensibility and Prehension Test, SIS- Stroke Impairment Scale, GAS- Goal Assessment Scale, ROM- range of motion, EMG- Electromyography, fMRI- Functional Magnetic Resonance Imaging*

**Table 3:** Relevant data extracted from studies for the eight studies included in the meta-analysis.

| Studies | Disease stage | No. of subjects | Outcome measure | Post (immediate) intervention FMA score change ($\mu \pm SD$) | SMD with 95% confidence interval | SE of SMD |
| --- | --- | --- | --- | --- | --- | --- |
| <sup>29</sup>Takahashi et al., 2008 | Chronic | EG: 7<br>CG: 6 | FMA | EG: $9.1 \pm 2.1$<br>CG: $5.8 \pm 1.6$ | 1.625 (0.983, 2.265) | 0.675 |
| <sup>28</sup>Ramos-Murguialday et al., 2013 | Chronic | EG: 16<br>CG: 14 | Modified FMA | EG: $3.4 \pm 0.563$<br>CG: $0.35 \pm 2.24$ | 1.877 (1.45, 2.303) | 0.448 |
| <sup>31</sup>Susanto et al., 2015 | Chronic | EG: 9<br>CG: 10 | FMA | EG: $5.11 \pm 6.55$<br>CG: $5.7 \pm 4.35$ | -0.103 (-0.539, 0.334) | 0.459 |
| <sup>36</sup>Frolov et al., 2017 | Subacute + Chronic | EG: 55<br>CG: 19 | FMA | EG: $4.67 \pm 17.83$<br>CG: $2.66 \pm 22.53$ | 0.104 (-0.149, 0.357) | 0.266 |
| | Chronic | EG: 30<br>CG: 10 | FMA | EG: $7.17 \pm 18.21$<br>CG: $4.0 \pm 24.33$ | 0.157 (-0.191, 0.504) | 0.365 |
| <sup>32</sup>Rowe et al., 2017 | Chronic | EG: 15<br>CG: 15 | FMA | EG: $2.7 \pm 2.6$<br>CG: $2.2 \pm 3.5$ | 0.157 (-0.189, 0.505) | 0.366 |
| <sup>22</sup>Wright et al., 2018 | Chronic | EG: 10<br>CG: 9 | FMA | EG: $1.1 \pm 1.69$<br>CG: $1 \pm 1.89$ | 0.053 (-0.383, 0.49) | 0.459 |
| <sup>27</sup>Cho and Song 2019 | Chronic | EG: 19<br>CG: 19 | FMA | EG: $4.0 \pm 2.58$<br>CG: $2.0 \pm 2.60$ | 0.755 (0.435, 1.076) | 0.337 |
| <sup>24</sup>Park et al., 2020 | Subacute + Chronic | EG: 10<br>CG: 9 | FMA | EG: $2.58 \pm 8.6$<br>CG: $5.4 \pm 7.48$<br>(data extracted from graph using web-plot digitizer) | -0.333 (-0.773, 0.107) | 0.463 |

\*EG- experimental group, CG- control group, FMA- Fugl-Meyer Assessment of the upper extremity, $\mu$ - mean, SD- standard deviation, SMD- standardized mean difference, CI- confidence interval, SE- standard error.
**Note:** The total score for FMA is 66, and for Modified FMA is 54 (the same outcome measure across studies but a different scale was used in one study; therefore, SMD was considered instead of mean difference (MD)).

For meta-analyses, upper-limb impairment was selected as the primary outcome of interest, typically quantified using the Fugl-Meyer Assessment (FMA) score. FMA scores were pooled from eight out of twelve included studies (as per availability), all involving people with stroke. Among these, seven studies provided numerical values of the outcome measure, while for one study^24^, values were extracted using WebPlotDigitizer^25^ from a plot showing FMA scores (Figure 4c of that study^24^). The change in FMA scores (pre- to post-intervention) representing the intervention effect was mined for both the experimental and control groups as mean ± standard deviation (Table 2). The standardized mean difference (SMD – the effect size) between the intervention outcome of experimental and control group was computed for individual study. Because all the included studies except one had small sample sizes (< 50), Hedges’ *g* SMDs (which corrects the sample bias unlike Cohen’s *d*) with its 95% confidence intervals were calculated and pooled across the studies. Based on Cohen’s criteria^26^, summary effect sizes were interpreted as small (0.2 – 0.49), moderate (0.50 – 0.79), or large (≥ 0.80). Both fixed-effect and random-effect models were utilized along with heterogeneity analysis. Heterogeneity was quantified using the estimated between-study variance (τ^2^) and percentage of variability attributed to heterogeneity (I^2^). When the I^2^ > 50% (moderate level), the summary effect obtained by applying the random-effects model was considered, as it accounts for between-study variance, providing a more reliable summary effect size. All analysis was implemented in Python (code and formulas provided in Supplementary Section III). The statistical significance was set at α = 0.05.

Meta-analysis was performed on the pooled data of all 8 studies to evaluate the overall intervention effect (i.e., effect of robotic assistance) irrespective of disease stage, the presence or absence of robotic assistance, the amount of assistance provided, and the targeted limb area. However, multiple subgroup analyses were performed on separate subsets of studies to determine the specific effect of robotic rehabilitation on chronic stroke survivors, and to compare the intervention effect for therapy targeting shoulder-elbow versus therapy targeting hand (wrist or fingers). In addition, sensitivity analysis was also performed for the overall effect; details of it are provided in Section-IV of the supplementary document.

## Results

Total 527 articles were screened, out of which 44 articles were assessed for eligibility. Upon thorough evaluation by the authors RC and SB, finally 12 articles (randomized controlled trials) were included for this systematic review and meta-analysis.

### Descriptive summary

A qualitative analysis of twelve controlled studies with sample sizes ranging from 13 to 74 (total sample - 339), examined the impact of robot-assisted upper-limb rehabilitation in people with neurological, including eight studies on stroke and two on other neurological disorders (spinal cord injury and multiple sclerosis). Six of the stroke studies were in chronic stroke, with other two covering a combination of subacute and chronic phases of stroke. The therapy dose varied from 4.5 to 28 hours, spanning 3 to 30 sessions over 2 to 8 weeks. The combined findings demonstrated a reduction in post-therapy impairment in both control and experimental groups. However, a few studies (25% of total studies) have shown a significant greater improvement in experimental group, indicating greater benefit could be obtained with assist-as-needed than constant assistance^27^, assistance contingent on movement intention than random assistance^28^, and complete active-assistance than a combination of active and no assistance^29^.

A greater functional and impairment recovery has been seen in moderately affected chronic stroke survivors when they received active-robotic assistance throughout the training duration as compared to when they received an equal combination of active assistance and no assistance^29^. For both the groups, the therapy was administered targeting hand movements through repetitive grasp-release task, and it showed a significant time effect for each group along with a significant between group difference post-therapy. The complete active assistance showed greater benefit than combination of active and no assistance^29^. Robot-assisted therapy was found to be more effective than unassisted therapy in functional performance of multiple-sclerosis survivors as well, who received shoulder-elbow training through repetitive reaching task^30^. In contrast, active assistance versus unassisted robotic therapy did not show any significant group difference when moderately affected chronic stroke survivors received hand training through finger flexion-extension task^31^. Similarly, no significant difference in improvement was found between active-robotic assistance and non-robotic assistance groups (both with gravity compensation) when shoulder-elbow training was delivered to moderately affected subacute and chronic stroke survivors through virtual reality-based tasks^24^. Although both the groups showed improvements, a greater change was seen in the non-robotic group for movement smoothness, functional and impairment parameters, whereas active robotic group exhibited better improvement only for mean movement speed. Comparisons between high and low robotic assistance showed a significant reduction in impairment and better functional outcomes in both the groups when moderately affected chronic stroke survivors received hand training^32^. Although no significant between group differences were found, a trend of greater impairment recovery, higher lateral pinch strength, and boosted motivation were seen in high assistance group, particularly for the survivors who had comparatively severe motor deficit. In addition, assist-as-needed mode (up to 4N assistive force when movement stopped for 2 sec) demonstrated significantly greater impairment recovery and functional improvement than guidance mode (a constant corrective and assistive force up to 4N) for shoulder-elbow training in moderately affected chronic stroke survivors^27^. In the same context, no significant outcome difference was seen in chronic incomplete spinal cord injury patients receiving elbow-wrist therapy with assist-as-needed or fixed subject-triggered robotic control^33^. Similarly, when comparison was made between deficit force field training and null force-field training for shoulder-elbow rehabilitation in moderate to severe chronic stroke survivors, it did not yield significant difference^21^. However, one study^34^ on severely affected chronic stroke survivors has reported that velocity dependant force (negative viscosity) training could facilitate movement performance (elbow extension flexion). Interestingly, it has also been observed that even when no active assistance was provided by robot, as compared to full arm-weight support training, the progressive arm-weight support led to significantly greater improvements in movement velocity, muscle strength, and work area for target-reaching tasks in people with moderate-to-severe chronic stroke^35^. Furthermore, robotic assistance contingent on movement intention led to superior impairment recovery of arm when compared with non-contingent robotic assistance in severely affected chronic stroke survivors^28^. In contrast, another study with similar intervention protocol for hand training but through motor imagery tasks, showed improvement in both the groups with no significant differences between the groups^36^. The experimental group consistently exhibited a significant difference in pre-post-therapy outcomes, whether chronic and subacute data (severely affected) were analysed together or separately. But the control group showed a significant difference only when chronic and subacute data were analysed together.

Overall, this qualitative summary highlights the contrasting findings across studies, wherein, only a few studies have shown positive effect of robotic assistance, and the rest are equivocal. Also, the risk of bias analysis (Figure 2) revealed that 67% of the studies lacked double blinding, potentially compromising the validity of the results. Therefore, nothing can be confidently commented on the role of robotic-assistance in upper-limb neurorehabilitation with the currently available RCTs.

**Figure 2:**
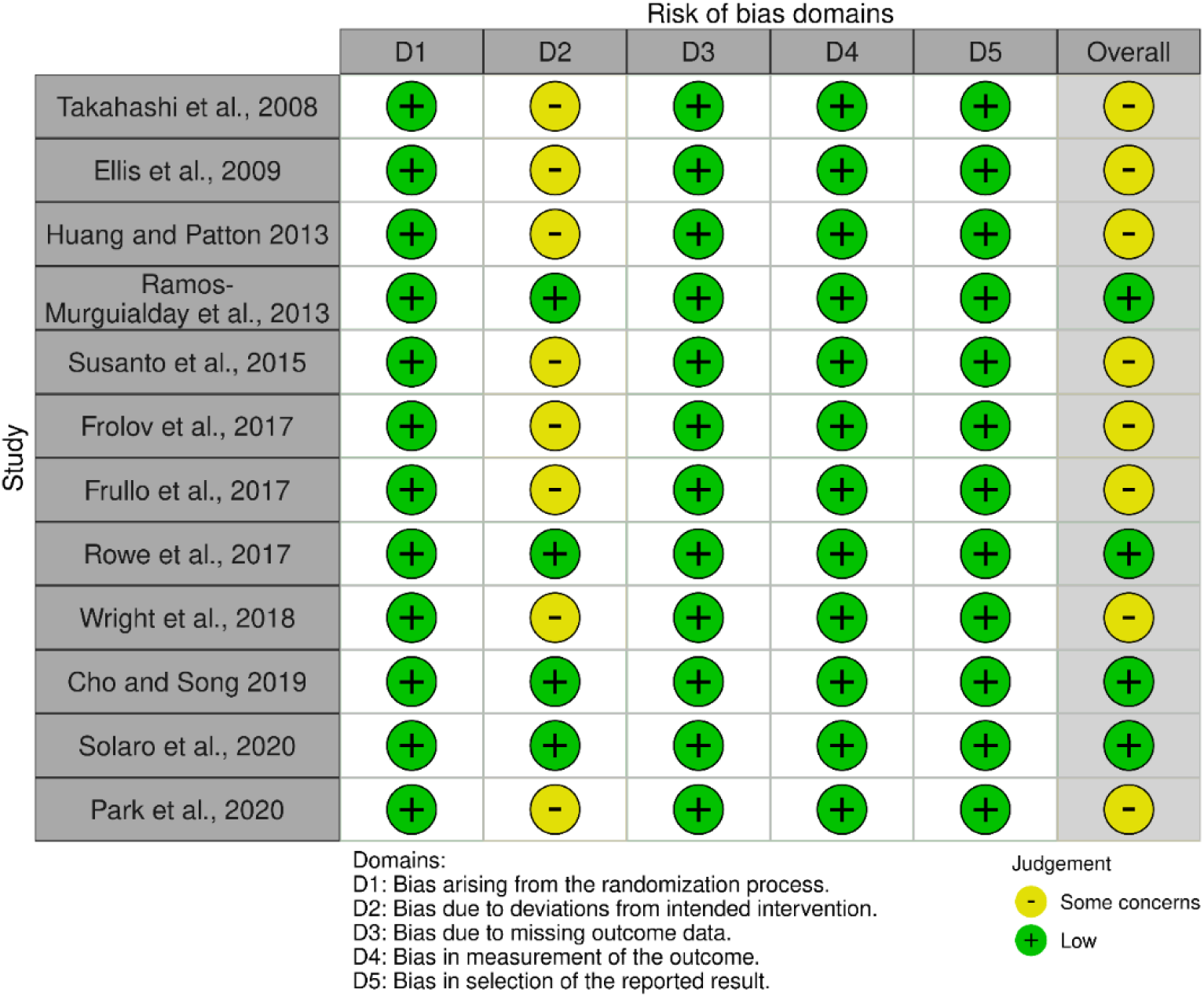
Risk of Bias analysis report

### Meta-Analysis

The main analysis was performed on the pre-post intervention FMA scores pooled from 8 controlled trials. The main analysis involved data of all 8 trials and subgroup analyses involved data of 3-7 trials based on specific aim of each analysis. No significant publication bias was found for these eight studies (see Section-IV of supplementary document). The consolidated outcomes of the main and subgroup meta-analyses, summarized using both fixed-effect and random-effect models, demonstrated effect sizes within the range of 0.201 to 0.588, indicating small to moderate effects of intervention. The corresponding heterogeneity analyses revealed 40.43 to 66.9% between-study variability (*I*^2^), with majority ≥ 50%, suggesting that the results should be interpreted with caution. Potential factors contributing to this substantial heterogeneity include inter-study differences in sample sizes, sample demographics, disease severity levels, robotic devices, movements/tasks trained, methodology, type of robotic assistance, and presence/absence of additional conventional therapy. Furthermore, as previously noted, the lack of double blinding poses a significant risk of bias.

A significant heterogeneous summary effect size (ES) was found for FMA score from the main meta-analysis involving 8 trials and 242 participants. The result exhibited a trend of favouring the experimental group (Figure 3(A); Table 4, row 2), indicating a positive impact of robot-assisted therapy in reduction of impairment regardless of the stage of the disease, mode of the assistance, level of the assistance and the targeted limb area.

**Figure 3:**
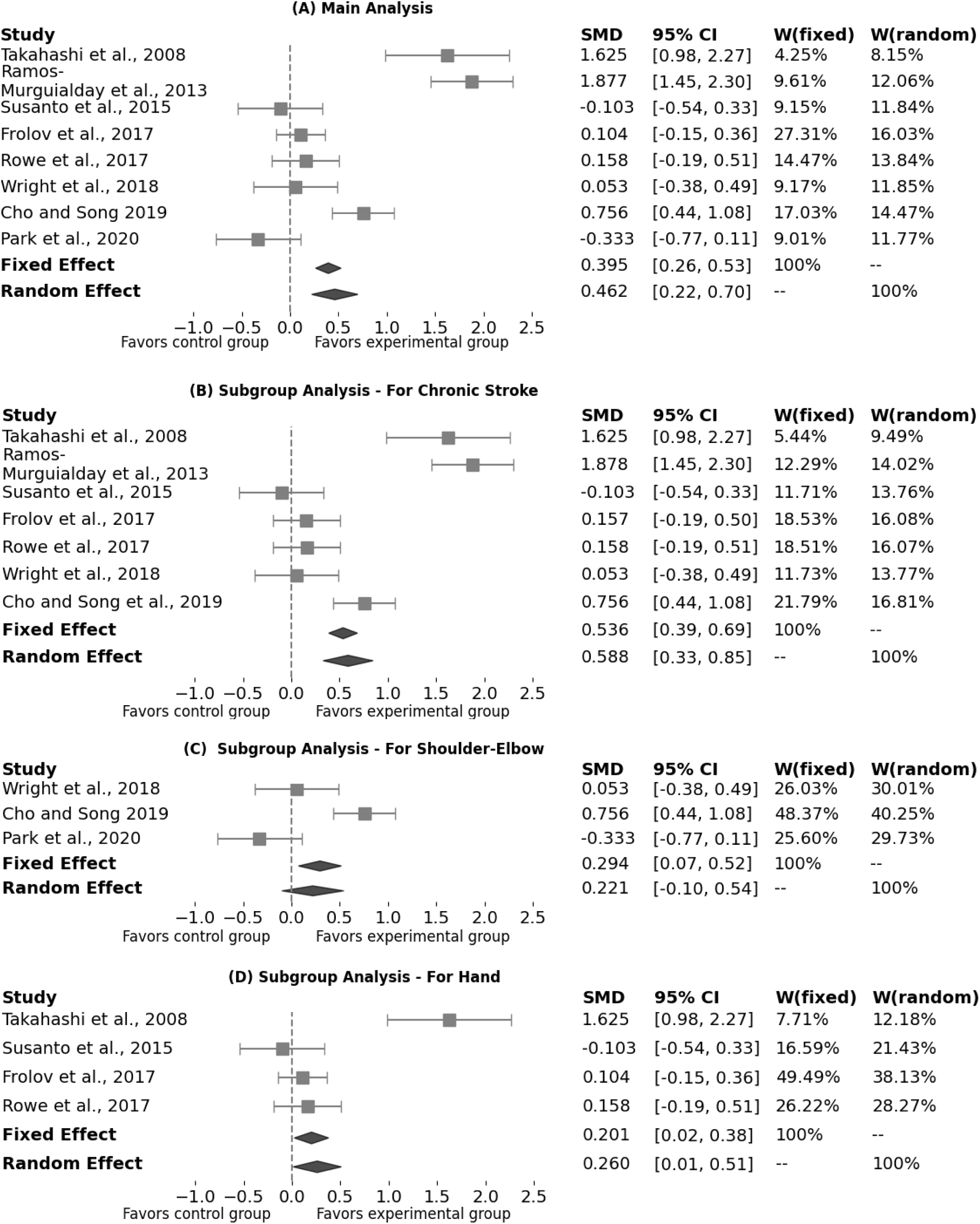
Forest plots representing multiple meta-analysis reports on FMA scores for (A) overall effect of robotic therapy, (B) chronic stroke, (C) therapy targeting shoulder-elbow, and (D) therapy targeting hand. For each study, the standardized mean difference (SMD) of FMA score with its 95% confidence interval (CI), and weights (W) for fixed-effect and random-effect models are given.

**Table 4:**
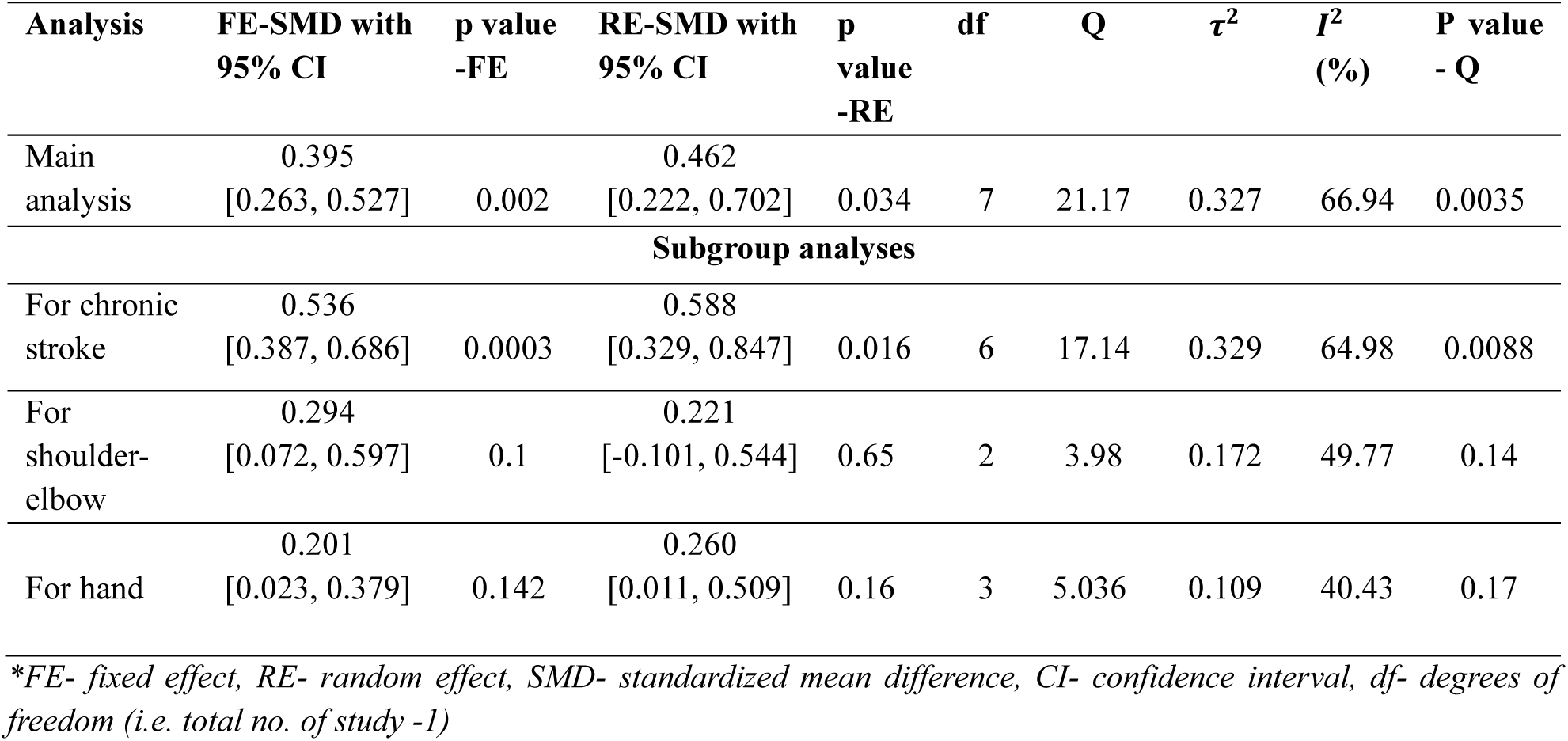
Meta-analyses reports.

| Analysis | FE-SMD with<br>95% CI | p value<br>-FE | RE-SMD with<br>95% CI | p<br>value<br>-RE | df | Q | $\tau^2$ | $I^2$<br>(%) | P value<br>- Q |
| --- | --- | --- | --- | --- | --- | --- | --- | --- | --- |
| Main<br>analysis | 0.395<br>[0.263, 0.527] | 0.002 | 0.462<br>[0.222, 0.702] | 0.034 | 7 | 21.17 | 0.327 | 66.94 | 0.0035 |
| **Subgroup analyses** |  |  |  |  |  |  |  |  |  |
| For chronic<br>stroke | 0.536<br>[0.387, 0.686] | 0.0003 | 0.588<br>[0.329, 0.847] | 0.016 | 6 | 17.14 | 0.329 | 64.98 | 0.0088 |
| For<br>shoulder-elbow | 0.294<br>[0.072, 0.597] | 0.1 | 0.221<br>[-0.101, 0.544] | 0.65 | 2 | 3.98 | 0.172 | 49.77 | 0.14 |
| For hand | 0.201<br>[0.023, 0.379] | 0.142 | 0.260<br>[0.011, 0.509] | 0.16 | 3 | 5.036 | 0.109 | 40.43 | 0.17 |

\*FE- fixed effect, RE- random effect, SMD- standardized mean difference, CI- confidence interval, df- degrees of freedom (i.e. total no. of study -1)

The subgroup analysis on studies (7 trials, 189 participants) with chronic stroke survivors showed a significant summary effect, indicating that robotic assistance could influence impairment recovery in chronic phase (Figure 3(B); Table 4, row 4). However, the significant heterogeneity observed in the analysis implies that further investigations are necessary to reach strong conclusions. The subgroup analyses for both shoulder-elbow training (3 trials, 76 participants) and hand training (4 trails; 136 participants) showed non-significant heterogeneous outcomes. However, the summary effect-sizes of these two cases are found to be similar, indicating that comparable amount of therapy gain could be obtained for shoulder-elbow and hand impairment recovery through robot-assisted training (Figure 3(C and D); Table 4, row 5 and 6).

## Discussion

The findings of this systematic review and meta-analysis highlight the potential beneficial effects of robotic assistance in upper-limb neurorehabilitation for people with neurological disorders, particularly stroke survivors. The qualitative synthesis of 12 RCTs and the quantitative analysis of 8 RCTs lend some support to the premise that robotic assistance improves motor recovery. However, no strong conclusions can be made from the existing literature due to the limited number of studies, their small sample sizes and high heterogeneity. The results from the current study also underscore the complexities of determining the optimal level and mode of robotic assistance or physical support, given the variability in patient responses and intervention parameters.

### Replicability of the included studies

A crucial aspect of clinical research is the ability to replicate findings across different settings and patient populations. While this meta-analysis aggregated data from multiple studies, variations in intervention protocols posed challenges to direct comparability and contributed to high heterogeneity across the studies. Many of the included trials lacked detailed information on key intervention parameters, such as robotic control algorithms, nature, content, and schedule of feedback, task/movement requirement and blinding procedures (details of the missing information from the different studies are provided in Section-V of the Supplementary Material). This lack of transparency limits the ability to reproduce findings in future research and undermines the reliability of the results. To avoid these issues, future studies should adhere to comprehensive and standardized reporting guidelines, such as the Consolidated Standards of Reporting Trials (CONSORT) checklist for randomized trials^37^. Implementing these guidelines will enhance rigor and improve the reproducibility of findings across diverse patient populations. Employing recent frameworks like the rehabilitation treatment specification system (RTSS) for detailed reporting therapeutic interventions will allow better synthesis of results from across studies^38^. In the context of robots assisted therapy, a similar framework for reporting the details of therapy implemented with the robot could be adopted. Standardized reporting of parameters related to robotic assistance, movements/tasks trained, and the feedback, can help better characterize inter-study differences, and thus evaluate the quality of results from systematic reviews and meta-analysis.

### Role of challenge and assistance levels

The concept of challenge in motor rehabilitation is crucial, as the appropriate level of difficulty can determine the effectiveness of therapy. High assistance may facilitate movement completion, adherence and improving motivation, especially in individual with severe impairments^31^. However, when assistance is too high, it may limit the engagement of intrinsic motor control mechanisms, reducing opportunities for voluntary effort (slacking)^39^ and skill acquisition. Conversely, assist-as-needed, resistive training and error-augmented training require patients to exert greater effort, which can enhance motor learning and neuroplasticity through increased proprioceptive and sensorimotor feedback. Studies have shown greater outcome of intervention for resistive training than assistive training^15,16,17^, active training than complete passive training^13,14^, and error-augmented training than error-reduction training^40^. These findings support the idea that increasing task challenge and active participation better engage the motor learning mechanisms and drive recovery more effectively than reducing difficulty. However, if the level of challenge exceeds the individual’s capabilities, it may lead to compensatory movements, or even disengagement from therapy. Studies indicate that the optimal approach is an adaptive assistance strategy, where robotic support is continuously adjusted based on the patient’s performance^2,41,42,43^. This ensures that the challenge remains within an optimal difficulty range, maximizing engagement and motor learning while preventing major dependency on robotic support^44^. Additionally, the impact of challenge level may vary based on impairment severity and the rehabilitation phase. For instance, in early rehabilitation, higher assistance might be necessary to initiate movement and prevent compensatory pattern^45-^. As recovery progresses, gradually reducing assistance and increasing task difficulty can encourage active participation and autonomous control of movements. Future research should focus on refining adaptive assistance algorithms that tailor challenge levels dynamically, ensuring optimal engagement and recovery across different patient profiles.

### Mechanisms underlying recovery through robot-assisted therapy

There are a few potential mechanisms through which robotic assistance might be beneficial, on top of benefits obtained from intense training. Robotic assistance might influence sensorimotor recovery by directly inducing cortical changes through Hebbian-like learning mechanisms^28,29,32,46,47^. Movement following neurological injuries are often diminished, slow, and poorly coordinated due to weak and altered motor output from the nervous system. Thus, the resulting afferent feedback is likely to be weak. Given the importance of afferent feedback in motor control and learning, it is reasonable to ask if stronger sensory feedback generated via repetitive, faster and larger movements through robotic assistance might be beneficial. The temporal alignment of this enhanced sensory input with movement intention is hypothesized to strengthen cortical and subcortical connections^48,49^ through mechanism such as spike-time dependent plasticity (STDP)^46,50^. However, how STDP functions at a circuit level, where pre- and post-synaptic activity occurs across diverse neural networks, remains unclear. In addition to cortical reorganization, robotic assistance may provide benefits at the musculoskeletal level. By enabling movement beyond the patient’s active range of motion, robots help stretch antagonist muscles and joint structures, potentially maintaining soft tissue integrity and preventing contractures. While stretching may temporarily reduce muscle spasticity, a systematic review and meta-analysis suggests that short-term protocols (e.g., three months) do not significantly alter joint range of motion^51^.

### Comparison with neuromuscular electrical stimulation (NMES)

Robot-assisted therapy is not the only form of assisted neurorehabilitation. Neuromuscular electrical stimulation (NMES) shares several traits with robot assisted neurorehabilitation, where external electrical currents are employed to activate intact peripheral neuromuscular structure to assist or generate limb movements^52,53^. NMES assisted movements are arguably more natural than robot-assisted movements, as the latter are generated through external forces. NMES not only induces agonist muscle contractions but also recruits several sensory structures during stimulation. Thus, arguably, NMES produces stronger afferent feedback than robots through (a) increased agonist muscle contraction, and (b) recruiting various sensory structures along the path of the stimulating current. Interestingly, peripheral motor nerve stimulation can also generate antidromic activity of the agonists, that can strengthen local spinal cord circuits through Hebbian learning^48^. Systematic reviews and meta-analyses provide some support to the benefits of NMES when combined with voluntary effort. Eraifej et al.^54^ reported a small improvement in upper limb impairments (measured by FMA) when NMES is combined with movement intent early on after stroke. Howlett et al.^55^ reported improved upper-limb activity with NMES compared to unassisted training. Both these reviews used sham or no stimulation during voluntary practice as their control groups. Bai et al.^56^ compared brain computer interface (BCI) triggered robot-assisted and NMES-assisted movements, and reported that BCI+NMES had significantly large improvements in upper limb functions, while BCI+Robot did not show any significant improvements, compared to sham BCI training or usual care. These systematic reviews support the tentative role of enhanced movement-related sensory feedback in promoting sensorimotor recovery, which warrant further investigation.

### Limitations

The current study has some limitations which must be considered when interpreting its outcomes. The current study focused only on RCTs involving groups differing in terms of the presence (assisted versus unassisted), level (low versus high assistance) or mode (fixed versus assist-as-needed control) of robotic assistance, and were specific to upper-limb rehabilitation. The outcomes do not say anything about the numerous other forms of human-robot physical interactions that have been implemented and tested with robots. These include, assistive versus resistive training, error-augmenting versus error-reduction training, active versus passive training, etc. Also, studies involving lower-limb or whole-body rehabilitation were not considered. This study did not examine the broader impact of robotic assistance on functional recovery or activities of daily living. The limited number of studies and the total number of participants included in the meta-analysis preclude any strong statements to be made about the effect of robotic assistance. However, the current work highlights the need for such studies to understand the mechanistic role of robotic assistance in recovery.

## Conclusion

This study investigated the role of robotic assistance on sensorimotor improvements through a systematic review and meta-analysis of RCTs on upper-limb robotic rehabilitation. The study provides preliminary evidence that the presence of robotic assistance might lead to better outcomes in upper limb impairments following neurological disorders, in particular stroke, compared to unassisted training in the same context. However, the limited number of appropriately designed studies (risk of bias), their tiny sample sizes, substantial heterogeneity, and the small values of the lower limits of 95% confidence interval of SMDs (≤ 0.3, indicating low effect sizes) limit any strong conclusion from the current literature. This study does highlight the dearth of rigorous, sufficiently powered studies evaluating the clinical value of the physical assistance from a robot during therapy. An enormous number of resources have been spent on the development of a diverse set of robotic devices (more than 38 types)^4^ for upper limb neurorehabilitation. Yet, we know shockingly little about the most fundamental and distinguishing feature of robot-assisted therapy – robotic assistance. De Iaco et al.^4^ call for an increased emphasis on the mechanistic understanding of robot-assisted therapy in their recent meta-analysis. This is essentially a call for the mechanistic understanding of the role of robotic assistance, because without physical assistance a robot is just a sensor. Thus, there is an urgent need of systematic, well-designed studies to evaluate the true clinical value of robotic assistance, if there is any.

## Data Availability

All data produced in the present work are contained in the manuscript

