## Supplementary Document for "Effect of Role of Robotic Assistance on Upper-limb Sensorimotor Recovery: A Systematic Review and Meta-Analysis"

**Section-I:**

Followings are the search strategies that were used for finding relevant papers in electronic databases

a) For advance search in PubMed-

**1.** ((Robot) OR (Robot Assisted) OR (Robotic Device) OR (End-Effector) OR (Exoskeleton) OR (Orthoses)) AND ((Active Assist) OR (Active Resist) OR (High Assistance) OR (Low Assistance) OR (Active Assistance) OR (Passive Assistance) OR (Unassisted) OR (Not Assisted)) AND ((Rehabilitation) OR (Neurorehabilitation) OR (Training) OR (Therapy) OR (Recovery)) AND ((Upper Limb) OR (Upper Extremity) OR (Hand) OR (Arm) OR (Shoulder-Elbow) OR (Wrist) OR (Finger)) AND (Control Group) AND (Patients) NOT (Review) NOT (Follow-up) NOT (Animal) NOT (Hip) NOT (Lower Limb) NOT (Leg) NOT (Knee) NOT (Foot) NOT (Gait) NOT (Surgery) NOT (Laparoscopy) NOT (Cancer)

**2.** ((Upper Limb) OR (Upper Extremity) OR (Hand) OR (Arm) OR (Shoulder-Elbow) OR (Wrist) OR (Finger)) AND ((Rehabilitation) OR (Training) OR (Therapy) OR (Recovery)) AND ((Robot) OR (Robotic Device) OR (Robot-assisted) OR (Robotic Assistance) OR (Robotic Training) OR (Robot-mediated) OR (Robot-based) OR (Robotic Rehabilitation) OR (End-effector) OR (Exoskeleton) OR (Orthoses)) AND ((Active Assisted) OR (Resistive) OR (High Assistance) OR (Low Assistance) OR (Active) OR (Passive)) AND (Control Group) NOT ((Review) OR (Animal) OR (Hip) OR (Lower Limb) OR (Leg) OR (Knee) OR (Foot) OR (Gait) OR (Walking) OR (Surgery) OR (Laparoscopy) OR (Cancer))

**3.** ((Robot-assisted) OR (Robotic Assistance) OR (Robotic Training) OR (Robot-mediated) OR (Robot-based) OR (Robotic Rehabilitation) OR (End-effector) OR (Exoskeleton) OR (Orthoses)) AND ((Active Assisted) OR (Resistive) OR (Assist-As-Needed) OR (High Assistance) OR (Low Assistance) OR (Active Assistance) OR (Passive Assistance)) AND ((Upper Limb) OR (Upper Extremity) OR (Hand) OR (Arm) OR (Shoulder-Elbow) OR (Wrist) OR (Finger)) AND ((Rehabilitation) OR (Training) OR (Motor Training) OR (Therapy) OR (Recovery) OR (Functional Recovery)) AND (Control Group) NOT ((Review) OR (Animal) OR (Hip) OR (Lower Limb) OR (Leg) OR (Knee) OR (Foot) OR (Gait) OR (Walking) OR (Surgery) OR (Laparoscopy) OR (Cancer))

**4.** ((Active Assisted) OR (Resistive) OR (Assist-As-Needed) OR (High Assistance) OR (Low Assistance) OR (Active) OR (Passive)) AND ((Robot) OR (Robotic device) OR (Robot-assisted) OR (Robotic assistance) OR (Robotic training) OR (Robot-mediated) OR (Robot-based) OR (Robotic Rehabilitation) OR (End-effector) OR (Exoskeleton)) AND ((Upper Limb) OR (Upper Extremity) OR (Hand) OR (Arm) OR (Shoulder-Elbow) OR (Wrist) OR (Finger)) AND ((Rehabilitation) OR (Training) OR (Motor Training) OR (Therapy) OR (Recovery) OR (Functional Recovery)) AND (Control Group) NOT ((Review) OR (Animal) OR (Hip) OR (Lower Limb) OR (Leg) OR (Knee) OR (Foot) OR (Gait) OR (Walking) OR (Surgery) OR (Laparoscopy) OR (Cancer))

b) For advance search in Scopus-

**1.** TITLE-ABS-KEY (("robo*" OR exoskeleton OR orthoses OR end-effector) AND (rehabilitation OR therapy OR training OR recovery) AND "assis*" AND ("upper limb" OR "upper extremity" OR hand OR arm OR "shoulder-elbow" OR wrist OR finger) AND "control group" AND NOT (review OR animal OR "lower limb" OR leg OR knee OR foot OR gait OR surgery OR laparoscopy OR cancer OR "follow-up"))

**2.** TITLE-ABS-KEY((“robo*” OR “exoskeleton” OR “orthoses” OR “brain-computer interface” OR “brain-machine interface”) AND (“assis*” OR “resis*”) AND (“upper limb" OR “upper extremity" OR “upper arm” OR hand OR arm OR “shoulder-elbow" OR wrist OR finger) AND (rehabilitation OR training OR “motor training" OR therapy OR recovery OR “functional recovery") AND (“control group”) AND NOT (review OR animal OR hip OR “lower limb" OR leg OR knee OR foot OR gait OR walking OR surgery OR laparoscopy OR cancer))

c) For advance and manual search in Google Scholar following keywords and multiple boolean combinations were used-

Google Scholar search- Robot, robotic device, robot assisted, exoskeleton, orthoses, assist, assistance, assistive, active assistance, passive assistance, high assistance, low assistance, unassisted, no assistance, resist, resistance, resistive, upper limb, upper extremity, hand, arm, wrist, finger, rehabilitation, neurorehabilitation, training, therapy

**Section-II**

Graphical representations of the information extracted from the selected papers

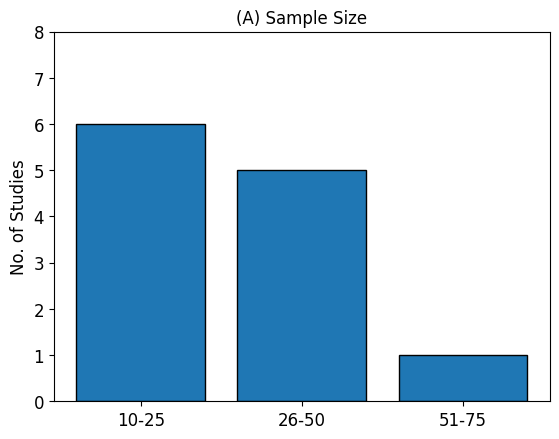

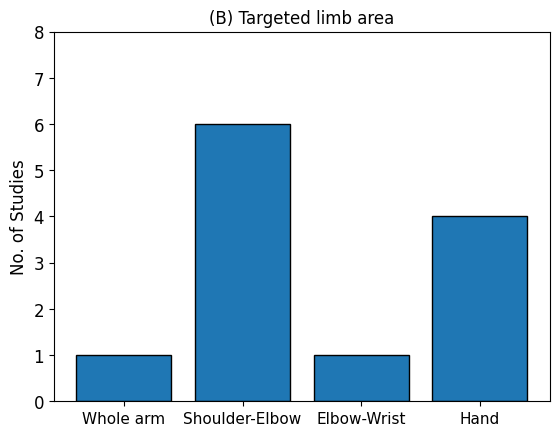

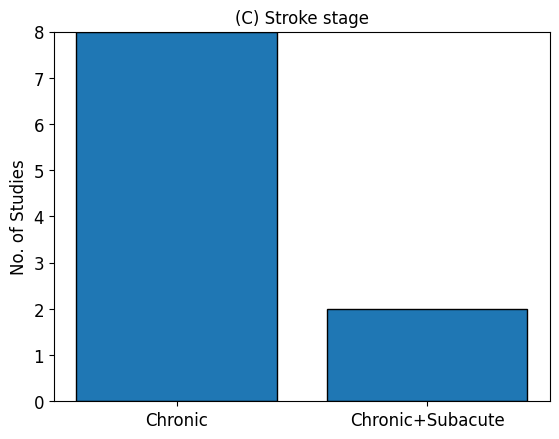

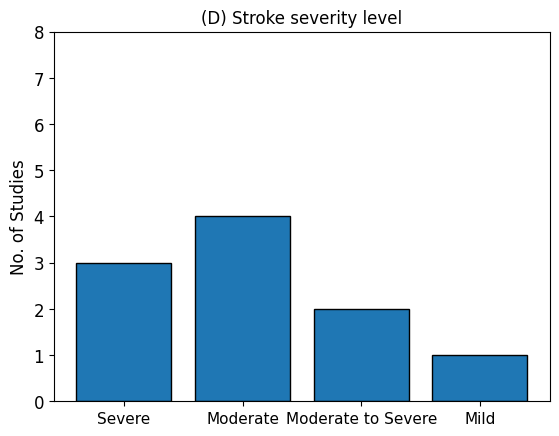

Figure 1: Histogram plots for (A) sample size, (B) targeted limb area, (C) stroke stage and (D) stroke severity level across studies

**Section-III**

Followings are the standard formulas that were used to compute the descriptive and inferential statistical parameters required for meta-analysis.

Across studies the data (Fugl-Meyer Assessment (FMA) score – primary outcome measure) were extracted in the form of mean ± standard deviation. When data were unavailable in the desired format, the change in FMA scores was calculated using pre- and post-intervention values, following the method described by Yagiz et al.^1^

Change in mean$(\Delta\mu)= \mu_{post}- \mu_{pre}$

Change in standard deviation ($\Delta SD)$ = $\sqrt{{SD}_{pre}^{2}+ {SD}_{post}^{2}- 2*r*{SD}_{pre}*{SD}_{post}}$

Here, $r$ is the correlation coefficient between the pre- and post-measurements; in none of the studies, the ‘$r$’ value was reported, so as per convention^1^, it was considered as 0.7. The main analysis was also repeated for two different values of ‘$r$’ (0.5 and 0.9) to evaluate the robustness of the study outcomes to changes in the value of ‘$r$’ (results are provided in the subsequent section i.e., Section-IV).

For studies that reported the (a) standard error (SE), or, (b) 95% confidence interval (CI), or, (c) interquartile range instead of SD, the corresponding SD was computed as follows^2,3^ –

$\left( a \right)SD= SE*\sqrt{no. of samples}$ ; or, $\left( b \right) SD= \sqrt{no. of samples}*\frac{(Upper limit-lower limit)}{4.128}$; or,

$$\left( c \right) SD=\frac{75\% quartile value-25\% quartile value}{ƞ}$$

Here, $ƞ$ is a dividing factor which is a function of sample size, for sample size > 50, $ƞ$ value was considered as 1.35, and for sample size ≤ 50, $ƞ$ value varied^3^ between 1.30-1.34.

Also, in a case, if median is reported instead of mean, then the mean was considered same as median for normally distributed sample, otherwise, the mean was computed as follows^3^ –

$$Mean=\frac{75\% quartile value+median+ 25\% quartile value}{3}$$

To obtain the summary effect of the combined outcomes of all/ a set of studies, following formulas^4,5^ was used-

(i) $SMD \left( \theta\right)= \frac{\mu_{E} - \mu_{C}}{\sigma_{pooled}}*j ; \sigma_{pooled}=\sqrt{\frac{\left( n_{E} -1 \right)\sigma_{E}^{2}+\left( n_{C} -1 \right)\sigma_{C}^{2}}{n_{E}+ n_{C} -2}} ;j=1-(\frac{3}{4*\left( n_{E}+n_{C}-2 \right)-1})$

Where, $SMD\left( \theta\right)$is the standardized mean difference, $\mu_{E}$ is the mean Fugl-Meyer Assessment (FMA) score change (pre to post intervention) of experimental group, $\mu_{c}$ is the mean FMA score change of control group, $\sigma$ is the standard deviation, $n_{E}$ is the no. of samples in experimental group, $n_{C}$ is the no. of samples in control group, $j$ is the correction factor for sample bias

(ii) ${SE}_{\theta}=\sqrt{\frac{n_{E}+n_{C}}{n_{E}n_{C}}+\frac{\theta^{2}}{2\left( n_{E}+n_{C} -3.94 \right)}} ; {CI}_{\theta}= \theta+(0.95*{SE}_{\theta})$

Here, ${SE}_{\theta}$ and ${CI}_{\theta}$ represent the standard error and confidence interval of SMD respectively

(iii) $W_{i(FE)}= \frac{1}{{SE}_{\theta}^{2}}; W_{Ni (FE)}= \frac{100* W_{i(FE)}}{\sum_{1}^{n} W_{i(FE)}}$

Here, $W_{i(FE)}$ and $W_{Ni (FE)}$ are the weights and normalized weights of the studies for fixed-effect (FE) analysis (inverse-variance method has been used for weight calculation)

(iv) Summary effect-size (SMD) of fixed-effect analysis: $\theta_{FE}= \frac{\sum_{1}^{n} {(W}_{i} * \theta_{i})}{\sum_{1}^{n} W_{i}}$

Here, $\theta_{i}$ and $W_{i(FE)}$are the SMD and weight of i^th^ study respectively; $n$is the total no. of studies

(v) Heterogeneity factors:

$$Q= \sum_{i=1}^{n} W_{i(FE)}\left( \theta_{i}-\theta_{FE} \right)^{2}; \tau^{2}= \frac{Q-df}{\sum_{1}^{n} W_{i(FE)} - \frac{{\sum_{1}^{n} W_{i(FE)}}^{2}}{\sum_{1}^{n} W_{i(FE)}}}; I^{2}= \frac{100 *(Q-df)}{Q}$$

Here, $df$ is the degrees of freedom = total no. of studies (n) - 1

(vi) $W_{i (RE)}=\frac{1}{{SE}_{\theta}^{2} + \tau^{2}}; W_{Ni(RE)}= \frac{100* W_{i(RE)}}{\sum_{1}^{n} W_{i(RE)}}$

Here, $W_{i(RE)}$ and $W_{Ni (RE)}$ are the weights and normalized weights of the studies respectively for random-effect (RE) analysis

(vii) Summary effect-size (SMD) of random-effect analysis: $\theta_{RE}= \frac{\sum_{1}^{n} (W_{i(RE)} * \theta_{i})}{\sum_{1}^{n} W_{i(RE)}}$

Statistical significance of the results was obtained using the Z-score^4^ for summary effects and Chi-square statistics for heterogeneity analysis. The respective formulas are as follows –

(viii) $Zscore= \frac{summary SMD}{SE of summary SMD}; SE of summary SMD= \frac{1}{\sqrt{\sum_{i=1}^{n} W_{i}}}$
(ix) $p value \left( one tailed test \right)=1- \emptyset(Zscore)$

where, $W_{i}$ is the weight of the *i^th^* study, $n =$ total number of studies, and $\emptyset$is the standard normal cumulative distribution function

(x) $p value \left( one tailed test \right)for heterogeneity=1- \chi^{2}(Q)$

where, $\chi^{2}$is the Chi-square distribution function and $Q$ is the Cochrane-Q value – a measure of between-study variability.

***For publication bias analysis:***

A scatter plot was obtained with effect size (as x value) and corresponding standard error (as y value) of individual studies. The pooled effect was calculated as the absolute mean of all the effect sizes. The following formulas^7^ are used for linear regression (Egger’s test) between precision (the inverse of standard error) and the standardized effect size, followed by intercept and p value computation.

$$\frac{\theta_{i}}{{SE}_{\theta_{i}}}= \beta_{0}+ \beta_{1}*\frac{1}{{SE}_{\theta_{i}}}+\epsilon_{i}$$

Where, $\theta_{i}$, ${SE}_{\theta_{i}}$, $\frac{\theta_{i}}{{SE}_{\theta_{i}}}$, $\frac{1}{{SE}_{\theta_{i}}}$are the effect size (standardized mean difference), standard error, standardized effect size and precision of the i^th^ study respectively; $\beta_{0}$, $\beta_{1}$ and $\epsilon_{i}$ are the intercept, slope and residual error of the linear regression fit respectively.

$p value \left( two tailed test \right)=2*(1- \emptyset(t)$)

Where, $\emptyset$is the standard normal cumulative distribution function, $t= \frac{\beta_{0}}{{SE}_{\beta_{0}}}$ (t statistics) for degrees of freedom (n-2); n is the total no. of studies.

***Link for data and python code -***

[*https://drive.google.com/drive/folders/1qi6-8ALz5OTooYSGrjuqGkR1SBK9RPXN?usp=sharing*](https://drive.google.com/drive/folders/1qi6-8ALz5OTooYSGrjuqGkR1SBK9RPXN?usp=sharing)

***Link for risk of bias (ROB) analysis document-***

[*https://docs.google.com/spreadsheets/d/1B243YBewx4PE0C5kHSxrFbsa_94shk28/edit?usp=sharing&ouid=114947611356048794179&rtpof=true&sd=true*](https://docs.google.com/spreadsheets/d/1B243YBewx4PE0C5kHSxrFbsa_94shk28/edit?usp=sharing&ouid=114947611356048794179&rtpof=true&sd=true)

Note: The ROB analysis was performed by answering the signaling questions by following the standard protocol^6^

**Section-IV**

a) Result of publication bias-

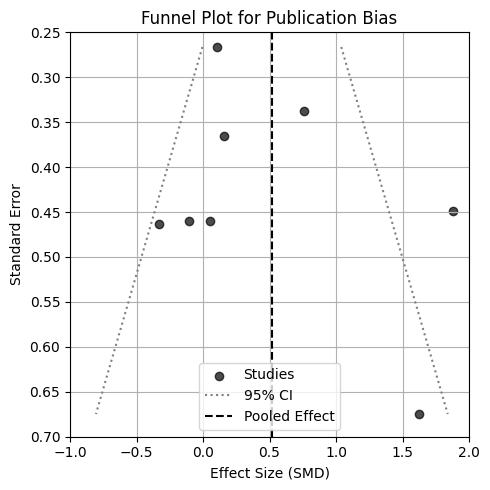

Figure 2: Funnel plot demonstrating the publication bias based on the effect sizes and standard errors of the eight studies that were included in the main meta-analysis. The black dotted vertical line is representing the abosulte mean of the effect sizes (pooled effect) and the grey dotted lines are representing the spread of its 95% confidence interval (CI).

**Observations:** Obtained results- pooled effect size = 0.517, intercept = 2.09 and p = 0.435; seven out eight studies are inside the funnel, indicating non-significant outcome; no significant assymetry, hence no significant publication bias has been found.

b) Sensitivity analysis-

Meta-analysis results (8 studies (df =7); sample size 242) when correlation coefficient (r) between the pre and post measurements was considered as 0.5, 0.7 and 0.9

| Main Analysis | FE-SMD with 95% CI | p value -FE | RE-SMD with 95% CI | p value -RE | Q | $\boldsymbol{\tau}^{\boldsymbol{2}}$ | $\boldsymbol{I}^{\boldsymbol{2}}$ (%) | p value - Q |
| --- | --- | --- | --- | --- | --- | --- | --- | --- |
| For  r = 0.5 | 0.372  [0.241, 0.503] | 0.0035 | 0.416  [0.205, 0.627] | 0.03 | 16.62 | 0.218 | 57.87 | 0.02 |
| For  r = 0.7 | 0.395  [0.263, 0.527] | 0.002 | 0.462  [0.222, 0.702] | 0.034 | 21.17 | 0.327 | 66.94 | 0.0035 |
| For  r = 0.9 | 0.418  [0.284, 0.553] | 0.0016 | 0.572  [0.261, 0.883] | 0.04 | 34.14 | 0.655 | 79.49 | 0.000016 |

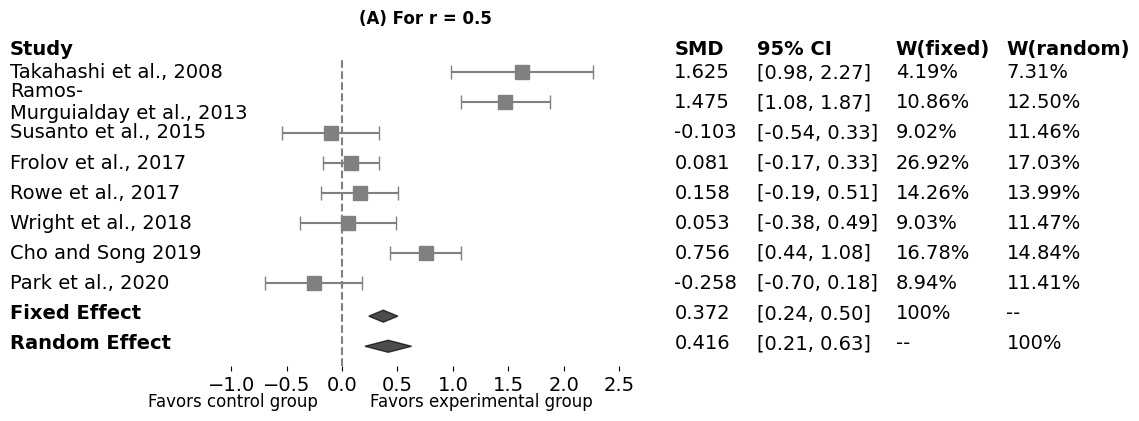

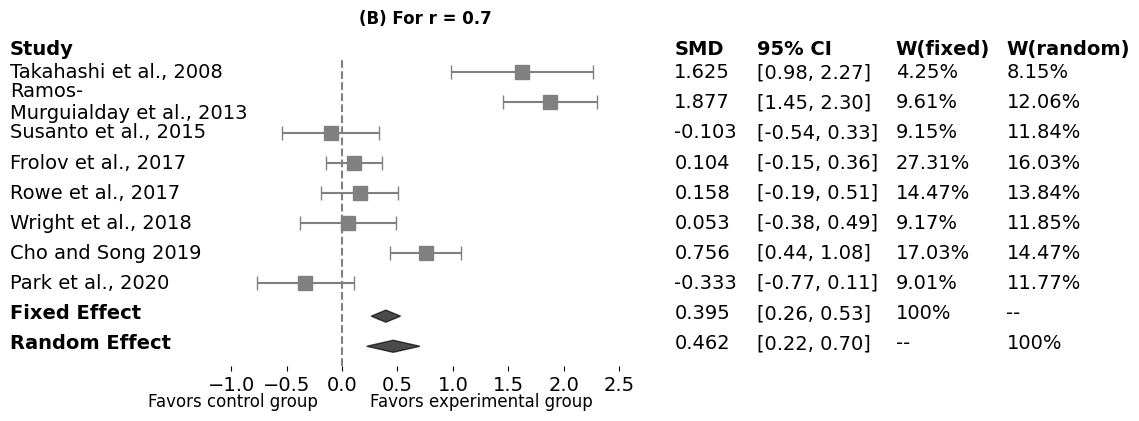

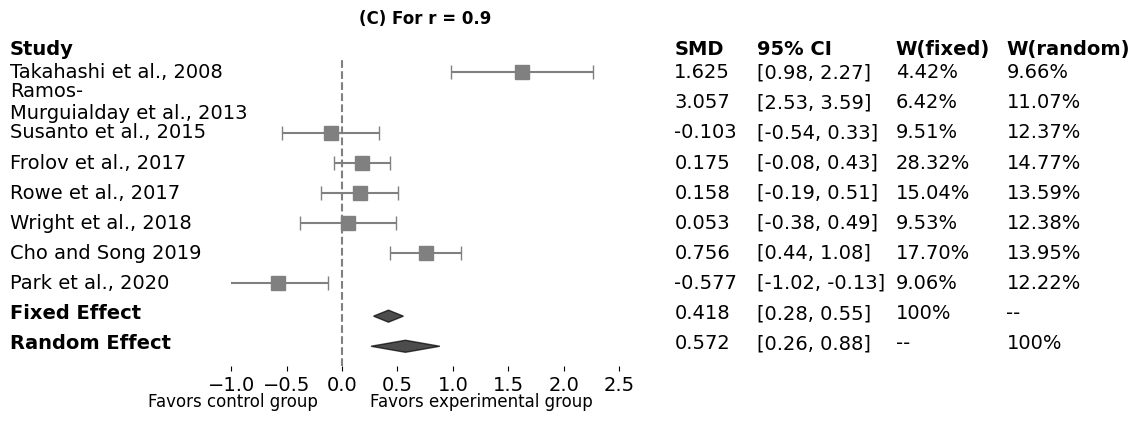

Figure 3: Forest plots for the robotic therapy meta-analysis on FMA scores when (A) r = 0.5, (B) r = 0.7, and (C) r = 0.9. For each study, the standardized mean difference (SMD) with its 95% confidence Interval (CI), and weights (W) for fixed-effect and random-effect models are given

**Observations:** Both fixed-effect and random-effect summary outcomes increase as ‘r’ increases, suggesting stronger observed effects when the correlation between pre- and post-measurements is higher**.** Higher correlation coefficients (r) result in larger effect sizes but introduce greater heterogeneity. Result for r = 0.9 showed the highest effect sizes and variability, whereas, r = 0.5 offered the most conservative estimates with relatively lower heterogeneity. However, this comparative analysis (sensitivity analysis) indicates that the direction of the obtained summary effect is stable for low to high pre-post measurement correlation.

c) Main meta-analysis report (9 studies; sample size - 256) that includes an addiditional study which apparently qualified the eligibility criteria but ultimately got excluded from core synthesis because of the ambiguity in its source reference, randomization and blinding procedure.

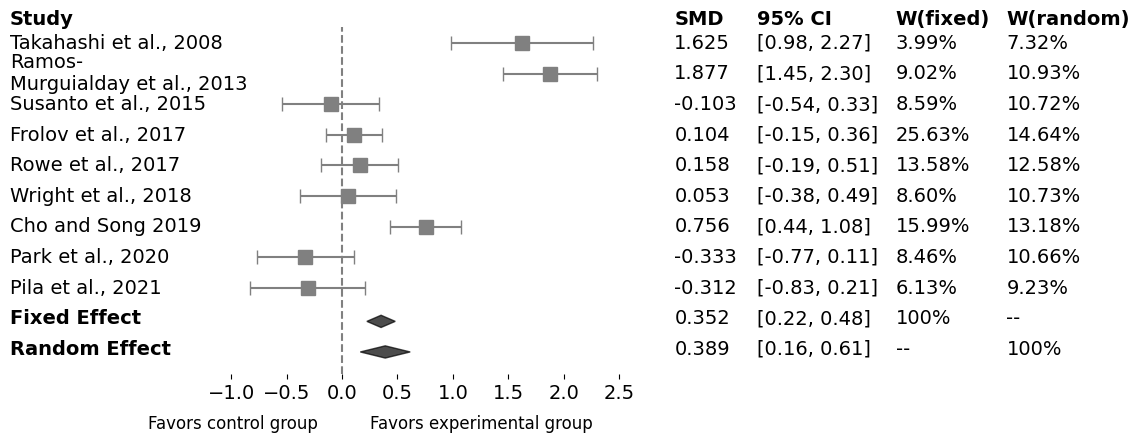

Figure 4: Forest plots for the robotic therapy meta-analysis on FMA scores. For each study, the standardized mean difference (SMD) with its 95% confidence Interval (CI), and weights (W) for fixed-effect and random-effect models are given

**Observations:** The obtained summary effect size is positive (fixed effect: 0.352 (0.224, 0.480); random effect: 0.389 (0.164, 0.615)), favouring the robotic assistance in imapirment recovery of upper limb irresective of disease stage, assistance level and targeted limb area. This is indicating a stable direction of main effect of robotic assistance with and without the additional study. As per the fixed-effect model, inclusion of this additional study has not remarkably altered the summary effect size (0.352 with p = 0.004) as compared to the summary effect size without the study (0.395 with p = 0.002). However, according to the random-effect model, inclusion of this additional study has caused a relatively greater shift in summary effect size (from 0.462 with p = 0.034 to 0.389 with p = 0.05) with a significant (p = 0.0037) heterogeinity (64.83%) across studies, making the result more inconclusive.

**Section-V**

Summary of the detials of the training provided by the studies chosen for the systematic review-

| **Study** | **Robotic Assistance** | **Movement target details** | **Movement characteristics emphasised** | **Movement details** | **Training feedback** | **Potential differences in the ingredients** | **Comments** |
| --- | --- | --- | --- | --- | --- | --- | --- |
| Takahashi et al., 2008 | **EG:** Full active assistance (A-A) throughout (15 days or 15 sessions)  Hard stop of the robotic assistance depended on patient’s passive ROM    **CG:** 7.5 days of training (half of the sessions) - active non-assist mode (ANA-A), but connected to the device, another 7.5 days of training - active assistance mode (A-A) | Grasp and release movement (11-15s long each).  For A-A: subjects were given 1-3s time to attempt to open or close fingers, after that assistance provided by the robot.  For ANA-A: subjects were given 3-5s time to attempt to open or close fingers    No information about the grasp object given | Game rule emphasized control of movement range, speed and timing. Accordingly, game difficulty levels were adjusted to avoid floor and ceiling effect    Also, task-specific motor cortex reorganization observed in fMRI analysis | 9 cycles of 10 repetition in first half of each day’s session, then15 min break, followed by second half of the session, wherein, subjects played VR based 9 games focusing grasp and release movement; A-A mode training with robotic assistance and ANA-A without assistance | Visual feedback given through LCD monitor, Audio instructions were given for grasp and release – ‘get ready’, ‘close’, ‘open’, ‘rest’. | First half of the total training days active participation was same between the groups, but in the second half of the days it was different | Study can be replicated from the details given about the intervention.  However, grasp strength and grasping object information are missing    Blinding information not mentioned clearly  In inclusion criteria, stroke onset time mentioned as at least > 3 months; mean and range of onset time are mentioned incorrect; it is confusing that included subjects all were chronic or mixed of subacute and chronic |
| Ellis et al. 2009 | **EG:** Progressive shoulder abduction loading  **CG:** Full shoulder abduction support  Robotic support but no assistance | Discrete planar reaching movements.  One target each in 5 different reaching directions.    No clear information about the location of the targets | Reach extent was emphasized; getting as close to the target is better. | 3 sets of 10 repetitions for each movement direction  No mention of the typical duration of each session.  30 second rests between repetitions, and 1 minute rest between sets. | Subjects received visual feedback from a computer monitor about trajectory of the actual movement.  Occasional/random feedback of movement performance (not clear what this feedback was about) was also given. No mentioned on the feedback schedule. | Active participation or effort was different between the groups due to the need for shoulder abduction in the experimental group. | Details about target locations is missing.  The robot does not provide active assistance for training; however, provided arm-weight support which aided the patients while performing the task.  Seeing change in FMA would have been interesting. |
| Huang and Patton 2013 | 3 sessions of training  **EG:** Two equal sub groups (cross over design)- Null force (no external force -N), negative viscous force (velocity dependent destabilizing force -V), and negative viscous force with inertia (Combined -C)  Condition randomization – NVC or NCV  **CG:** Null force filed (NNN) | **Training phase-** any movement of own choice using a variety of directions, speed and positions within a rectangular workspace (exploratory movement for 25 min)  **Evaluation phase-**four complete counter-clockwise revolutions around a target (circular tracking movements) | Speed, accuracy and smoothness of movement | 2 hours 15 min session duration. Each session included several alternating training phases (16) and evaluation trials (160). Experimental protocol not clear.  No clear information about movement repetitions | Real-time feedback of the handle position, visual reference cues, and experiment instructions were presented on a horizontal surface overlying the planar workspace of the arm. | Active participation or effort is likely to be different between the two groups because of the nature of the intervention | Study can be replicated from the details given about the intervention.    Blinding information not mentioned clearly  Groups were not balanced in terms of the severity level of the patients as the baseline FMA score for all the patients were not available/ measured |
| Ramos-Murguialday et al., 2013 | **EG:** robotic assistance contingent on movement intension (concurrent movement of orthoses)    **CG:** robotic assistance non-contingent on movement intension (random movement of orthoses) | The patients were instructed to move their arm and try to reach forward (even if the robotic arm orthosis does not follow their intention), then grasp, and bring an imaginary apple to their lap, thus involving finger extension during the reach and grasp movement. Each trail was of 5s.    After 8^th^ session, the BMI training was switched from arm to hand. | Not clear what aspects of the movement were emphasized | Approximately 13 – 15 min session duration (estimated by the reader; no distinct information provided by the authors). | No mention about feedback for task result or task performance | Active participation or effort was likely to be different between the two groups because of the nature of the assistance. | How the concurrent movements of the arm and hand orthoses took place that is not clear. Also, no information about reaching distance, grasp size.  Study may not be replicated with the information provided |
| Susanto et al., 2015 | **EG:** robot (exoskeleton) assisted    **CG:** attached to the exoskeleton but unassisted | Three modes of training- hand grasp-open (4 min), three-finger pinch-open (8 min), two-finger pinch-open (8 min); 1-2 min rest after each block. In all cases, subjects were instructed to move a kitchen sponge on a horizontal plane along a predefined direction within the predefined training space | Not clear what aspects of the movement were emphasized (speed, accuracy, quality)    Improvement in number of repetitions was one of the outcome measures | 1 hour session duration. Task related information is sufficiently provided | Verbal instructions and feedback were given; start and end points of movements were marked for subjects’ ease of doing the task | Active participation or effort was likely to be different between the two groups because of the nature of the assistance | The study can be replicated from the details given about the intervention.  The control group didn’t get assistance from robot but got it from therapist.  The result may be confounded due to the therapist assistance; role of robotic assistance is supressed.  Single blinded study |
| Frolov et al., 2017 | **EG:** Assistance dependent on motor imagery-based EEG activity  **CG:** Assistance non-dependent on motor imagery-based EEG activity; sham BCI | Subjects performed three mental tasks- motor relaxation, motor imagery of paretic hand opening, and motor imagery of non-paretic hand opening.  **BCI active-** the task-imagination specific brain activity drove the exoskeleton attached to paretic hand **BCI passive-** actuation of the exoskeleton based on the cue (appeared arrow on the screen), not based on the brain activity | Not clear what aspects of the movement were emphasized (speed, accuracy, quality) | 40 min session duration  Visual instructions on a computer screen were provided, wherein three coloured arrows indicated which mental task to be performed when. | The subjects received both visual and kinaesthetic feedback  of the results of BCI decoding of their imagery attempts | Active participation or effort was likely to be different between the two groups because of the nature of the assistance | The study can be replicated from the details given about the intervention. |
| Frullo et al., 2017 | **EG:** Assist as assisted with triggered assistance when movement stops    **CG:** Self-triggered control (partial passive assistance); full robotic assistance was provided when subject initiated movement crossed a threshold force towards intended direction | Subjects performed point to point reaching movement from centre position to peripheral target position (in the active ROM) | Movement velocity and number of repetitions | 90 min session duration. Not clear how much time was spent moving and resting | Subjects received visual feedback from a computer screen (home position, target position, desired trajectory, direction to which force should be applied) | Active participation or effort was likely to be different between the two groups because of the nature of the assistance. | The study can be replicated from the details provided about the intervention    Blinding information not mentioned clearly |
| Rowe et al., 2017 | **EG:** High assistance; (causing 80-85% success rate at hitting targets)  **CG:** Low assistance; (causing 50-55% success rate at hitting targets)  The amount of robotic assistance was varied between groups while number, amplitude and exerted effort of training movements were controlled. | Robot assisted the participants by applying guiding forces to direct their fingers along a physiological spatiotemporal trajectory, ensuring they reached the musical note at the target. However, these assistive forces were only activated when participants initiated the movement themselves, as detected by force sensors positioned between their fingers and the robot mechanisms, with a threshold set at 6 N. | Movement trajectory and force.  Movement trajectory intercepted the musical note at the target. | 1 hour session, moving index and middle finger to the targets to play a musical game. Total of 1065 possible movements per session | Subjects received visual feedback from a computer screen about the trajectory, moment of crossing and the angular distance to the actual crossing | Active participation or effort was likely to be different between the two groups because of the nature of the assistance. | The study can be replicated from the details provided about the intervention given to the subjects. |
| Wright et al., 2018 | **EG:** Deficit force-field robotic assistance    **CG:** Null force-field robotic assistance | **Training-** two blocks of eight, 2 min motor exploration trials (total 32 min)  During exploratory movement task, a trial was ended after two cumulative minutes of movement with speed > 0.04m/s; 1-3 min intermittent rest period  **Performance assessment**- 30 trials goal directed reaching task; 6 trials circular movement task; 6 trials - exploratory movement task | Movement trajectory and velocity | Training duration in a session- 33 to 35 min including rest.  Task related information is adequately given | Real-time visual feedback of the robot’s handle location  Additionally, randomness score for exploratory movement | Active participation or effort was likely to be different between the two groups because of the nature of the assistance. | Study can be replicated with the information provided about the intervention    Blinding information not mentioned clearly |
| Cho et. al 2019 | **EG:** Assist-as-needed mode - triggered assistance (maximum 4N assistive force); when movement stops for 2 sec, assistance is ON    **CG:** Guidance mode- corrective and assistive forces (maximum 4N) were applied during reaching to a straight-line path. Assistance was always ON. | Discrete 3D reaching movements; 6 different targets in 6 directions at desk and chest level  No mention of where the targets were located with respect to the subject; no information about reaching distance for individual target    Not clear how target locations were scaled to subject’s arm length | Not clear what aspects of the movement were emphasized (speed, accuracy, quality). | 40 min session duration. It is not clear how much time was spent moving and resting. No mention of the number of movement repetitions | Subjects received visual feedback from a computer screen about the trajectory of the actual movement.  Audio feedback on successful target reach. | Active participation or effort was likely to be different between the two groups because of the nature of the assistance. | The study cannot be replicated from the details provided about the intervention given to the subjects.  No clear mention about subjects’ severity. Not clear- Does “subjects were all high functioning” imply they were mildly affected? |
| Solaro et al., 2020 | **EG:** Haptic- robot assisted  **CG:** Sensorimotor- visual control- no force applied by the robot | Fast and accurate point to point (22.5 cm) reaching task in 6 different directions (0^o^, 60^o^, 120^o^, 180^o^, 240^o^, 360^o^) | Movement trajectory and reaching time | 40 min session duration. 8 epochs in session; in each epoch- 4 movement repetitions for each direction (thus, total 24 movements).  No information about rest time | Subjects received visual feedback from a computer screen.  Auditory feedback was also provided  after task completion (pleasant or unpleasant sounds depending  on the score). | Active participation or effort was likely to be different between the two groups because of the nature of the assistance and difference in task difficulty level | Although subjects of control and experimental group performed same task, the difficulty level was greater for experimental group    Study can be replicated from the details provided about the intervention |
| Park et al., 2020 | **EG:** Active robotic assistance with gravity compensation  **CG:** Unassisted; only mechanical support with gravity compensation by the robot; no actuation | Subjects were trained  with a game-based virtual reality environment with a  focus on proximal upper limb movement. | Not clear what aspects of the movement were emphasized (speed, accuracy, quality). | 30 min session. No information about the games, movement type, direction, number of repetitions and rest period | Subjects received visual feedback from a computer screen. | Active participation or effort was likely to be different between the two groups because of the nature of the assistance | The study can be replicated from the details provided about the intervention given to the subjects.    Blinding information not mentioned clearly |
